# Remote Sweat Chloride and Heart Rate Monitoring Reveal Variable Sweat Salt Loss During Exercise in Patients with Cystic Fibrosis

**DOI:** 10.64898/2026.07.06.26357386

**Authors:** Thaddeus Cybulski, Rachel S. Nelson, Margaret G. Grossman, Zasu M. Klug, Mia Calamari, Alvaro Donayre, Leah J. Welty, Susanna A. McColley, Jared Schooley, Garett J. Griffith, Daniel M. Corcos, Donald E. Wright, Jessica C. Wallace, Da Som Yang, John A. Wright, John A. Rogers, Roozbeh Ghaffari, AJ Aranyosi, Manu Jain

## Abstract

Cystic fibrosis (CF) is characterized by defective CFTR-mediated chloride transport, resulting in elevated sweat chloride concentrations. As people with CF (PwCF) now live longer due to highly effective CFTR modulators, exercise has become integral to maintaining health, yet it introduces additional physiological demands on salt and fluid balance. In this study, we used a wearable microfluidic biosensor (CF Patch) to quantify sweat rate and chloride loss during exercise performed both in the supervised laboratory and remote free-living in PwCF and healthy volunteers (HV). Participants completed exercise sessions under both conditions, with continuous heart rate monitoring and sweat collection with real-time measurement of sweat characteristics. Sweat volume and chloride concentration were assessed by colorimetric image analysis, enabling estimation of total fluid and chloride loss at the end of each exercise session. PwCF exercised for a longer duration at a lower average heart rate during remote exercise compared to laboratory exercise though exercise volume (average heart rate x duration) was greater during remote exercise. There was a positive association between exercise volume and both fluid and chloride loss for both PwCF and HV. PwCF exhibited greater chloride loss for a given exercise volume compared to HV, though fluid loss was similar. Further, compared to HV, PwCF demonstrated significantly greater intra- and interindividual variability in sweat chloride loss across the remote exercise sessions. Collectively, these findings provide evidence for the feasibility and physiological validity of remote exercise assessment and establish the feasibility and physiological validity of wearable sweat sensing for remote monitoring of fluid and electrolyte dynamics during real-world exercise. In addition, the variability of chloride loss in response to exercise suggests utility of the CF Patch in providing personalized fluid and salt repletion data for PwCF and advances the translational potential of digital sweat diagnostics for personalized CF care.

## Introduction

Cystic fibrosis (CF) is a life-shortening genetic disease caused by mutations in the CF Transmembrane Conductance Regulator (*CFTR*) gene, leading to impaired epithelial chloride transport and elevated sweat chloride and sodium concentrations. Measurement of sweat chloride remains a cornerstone of CF diagnosis and a robust biomarker of CFTR function. With the advent of highly effective CFTR modulators, this biomarker has the potential to evaluate therapeutic responses and disease trajectory. However, conventional pilocarpine iontophoresis and chloridometry are limited to specialized centers and predominantly pediatric hospitals, provide only static assessments of a highly dynamic physiologic parameter and are primarily used for CF diagnosis. This limitation restricts the ability to capture temporal variability in CFTR function, treatment adherence, or other factors that may significantly influence clinical outcomes in the era of modulator therapy.

CFTR modulators have profoundly altered the natural history of CF, extending life expectancy and improving healthspan for most people with CF (PwCF).(1) While it has long been known that increased aerobic capacity is associated with reduced mortality in CF,(2) exercise has assumed greater importance not only as a means of maintaining pulmonary health and muscle mass but also as a contributor to cardiovascular fitness, bone density, mental health, and overall well-being.(3–6) Exercise is increasingly prescribed as a standard adjunct to medical therapy, yet it introduces physiological stress that influences hydration, electrolyte balance, and sweat composition.(7) Understanding how CFTR dysfunction and modulator therapy affect salt loss during exercise is therefore clinically relevant—not only for optimizing hydration and electrolyte replacement but also for leveraging sweat-based biomarkers to monitor CFTR function under real-world physiological conditions.

Wearable microfluidic biosensors have emerged as a powerful platform for remote and serial physiological monitoring during exercise and other conditions.(8–11) The CF Patch, a skin-interfaced, colorimetric microfluidic device coupled with smartphone-based imaging, enables real-time quantification of sweat chloride and sodium concentrations during natural exercise.(12) Previous validation studies have shown strong agreement between CF Patch–derived sweat chloride and gold-standard pilocarpine chloridometry, and feasibility for serial remote use in both PwCF and healthy volunteers.(12) Importantly, PwCF on CFTR modulators demonstrated greater day-to-day variability in sweat chloride than healthy individuals, suggesting that real-world sweat monitoring could reveal fluctuations in CFTR activity or therapeutic responses that are not captured by single clinic-based measurements.

In this study, we extend these observations by quantifying exercise-induced salt loss in PwCF and healthy volunteers using the CF Patch. By characterizing both inter- and intra-individual variability across repeated exercise sessions, we aimed to investigate the physiological and therapeutic determinants of salt loss and assess the translational potential of wearable sweat sensing for personalized CF management. This work supports a new paradigm in CF care—where continuous, real-world physiologic data can inform adherence, CFTR modulator efficacy, and individualized recommendations for exercise, hydration, and electrolyte repletion in an aging CF population.

More broadly, this study contributes to the growing field of digital biomarker development and remote precision medicine in CF. By integrating wearable sensing, mobile analytics, and cloud-based data capture, the CF Patch represents a scalable framework for longitudinal monitoring of CFTR function beyond the clinic. Such platforms could enable detection of treatment non-adherence, point-of care monitoring, expanded assessment of modulator pharmacodynamics, and personalized feedback to optimize both therapy and lifestyle interventions. As CF care evolves toward proactive, data-driven disease management, wearable sweat monitoring offers a clinically meaningful bridge between molecular correction and real-world physiology—transforming how we track, interpret, and ultimately individualize therapy for people with CF.

## Methods

### Study design

We conducted a prospective, single-center, feasibility and exploratory study of adult PwCF and adult healthy volunteers (HV) who self-identified as exercising at home. The primary objective of the study was to establish feasibility of remote sweat and exercise monitoring in HV and PwCF populations, with secondary exploratory objectives of examining potential differences in exercise and sweat characteristics between the populations. The study was approved by the Northwestern University Biomedical Institutional Review Board (IRB) (STU00215214). Participants were recruited between April 2022 and May 2024. PwCF were recruited from a single CF center at Canning Thoracic Institute (Northwestern Medicine) and HV were recruited from the community. All participants were informed of the experimental procedures and associated risks before providing consent.

Each participant was asked to complete 2 parts of the study. Participants first completed clinic-based sweat testing, consisting of resting pilocarpine-induced sweat collection and a cardiopulmonary exercise test (CPET) with on-site exercise-induced sweat collection at the exercise physiology laboratory of the Department of Physical Therapy & Human Movement Sciences (Northwestern University). After completing the on-site portion, participants were asked to complete 5 remote exercise sessions over 14 days while wearing CF sweat patches. The location, mode, duration, and intensity of remote exercise were of their own preference.

### Human subject sweat and exercise testing

Each subject was asked to complete a symptom-limited CPET. CPET consisted of an incremental protocol on a cycle ergometer (LODE, The Netherlands). Once initiated, work rate increased incrementally every minute until the subject was no longer able to maintain a cycling cadence of ≥ 60 revolutions per minute. Rating of perceived exertion and exercising blood pressure values were recorded every two minutes during the CPET. Peak aerobic capacity (VO_2peak_) was determined using 30-second averaging, and the highest single 30-second value was recorded as VO_2peak_. Exercising heart rate data throughout the study were monitored by a Medtronic Zephyr™ biomodule and chest strap (Medtronic, Minneapolis, MN).

After completing the CPET, participants were allowed a dedicated rest period and fluid replenishment (water and Gatorade^TM^) until their baseline heart rate was reached and post-CPET sweat induction had ceased. Research personnel then applied two microfluidic sweat patches, one on each arm after which each subject began exercising at a constant wattage which was 50-70% of peak wattage derived from the CPET, i.e., submaximal-work exercise. Exercising heart rate and rating of perceived exertion, as well as environmental conditions including temperature, barometric pressure, and humidity of the exercise physiology laboratory were recorded every 5 minutes during on-site exercise. Participants continued exercising until either there was sufficient sweat in each channel of the CF Patch (assessed by visual inspection) or 60 minutes had elapsed.

### Remote exercise sessions and sweat and heart rate collection

Following recovery from exercise testing, participants were provided with CF sweat patches and a Medtronic Zephyr™ biomodule with chest strap to wear during the 5 remote exercise sessions over a 14-day period. Subjects were trained on use of all devices prior to starting remote workouts. They were also provided with a smart phone with the OmniSense™ mobile application. There were no specific requirements for the remote exercise sessions and participants were instructed to perform their normal exercise activities (e.g., running/cycling, resistance training, hot yoga) for their usual intensity and duration. Participants were instructed to take photos of the CF sweat patches following each of the 5 remote exercise sessions and transmit the images to the study team to be analyzed. Heart rate data were transmitted second-by-second to the OmniSense™ application from the Zephyr™ biomodule for the duration of each remote exercise session. Participants who did not have fully successful data collection in all 5 sessions were offered the opportunity to complete an additional home exercise session. If completed, this additional session was substituted for the exercise session with the most missing data (heart rate and/or sweat), with temporal proximity breaking ties. Three participants had substituted data. Exercise intensity was quantified by recording mean exercising heart rate, and volume of exercise was calculated as the product of mean exercising heart rate (bpm) and duration (min).

### Data collection

Study data were collected and managed using Research Electronic Data Capture (REDCap) tools hosted at Northwestern University.(13–15)

### Smartphone and camera-based colorimetric analysis of sweat volume and Cl concentration

Details on smartphone colorimetric analysis under controlled and ambient lighting conditions have been published.(11) Briefly, photos of microfluidic patches were captured using a smartphone application (iPhone 11, Apple Inc.) in clinical and remote settings. Images were captured at the time of CF Patch removal at the end of sweat-induction. When captured in a clinical setting multiple images were captured by study personnel within 5 minutes of completion of sweat collection. At home, study participants were instructed to capture multiple CF Patch images within 5 minutes of exercise completion. To assess feasibility of remote image capture, success was deemed if a participant recorded interpretable images from at least 2 remote exercise sessions. Final mean volume and sweat [Cl^-^] were calculated from manual image analysis. Sweat [Cl^-^] and volume data from CF Patch collections were obtained from each sample from which at least a single image measurement was available. RAW-format images were used for processing to eliminate artifacts introduced by normalization, compression, and other preprocessing steps. The sweat volume and [Cl^-^] were calculated by manual image analysis by a trained data scientist.(10) The colors of four swatches on the CF Patch (green, purple, orange, white) were measured and used to correct for variations in white balance due to ambient lighting. Local sweat volume was measured by identifying the furthest extent to which colored dye propagated in microchannel 1 and mapping that location to a volume using a 3D model of the patch. Chloride was measured by computing the average color within microchannel 2 relative to the background color of the patch. In CIELAB color space this color is represented by a 2D vector. This vector was then projected onto the unit vector for the color of chloranilic acid, which was measured empirically. The length of this projection maps to chloride concentration by a power law relationship, as described by the Beer-Lambert law.

### Statistical analysis

We used Stata version 17 (16)for descriptive and inferential statistics, and R version 4.4.1(17) for data visualization. Descriptive statistics are reported as mean (standard deviation) and frequency (percent). We compared onsite exercise characteristics between PwCF and HV using standardized mean differences because we are interested in the magnitude of group differences, rather than if differences are statistically significant and non-zero.

We used linear mixed effects models to estimate differences in sweat loss, chloride loss, and exercise characteristics between PwCF and HV and between onsite and remote exercise sessions within group. We included fixed effects for group (PwCF versus HV), and for models comparing onsite and remote exercise, an interaction between group and exercise location. We also report models for total chloride loss that adjust for exercise volume by including it as a fixed effect; adjustment is noted in the text. To determine appropriate random effects, we used likelihood ratio tests to compare nested models. Because we hypothesized that measurements on PwCF would be more variable than those on HV, we compared: (1) models with random intercepts for participants to models with random intercepts for participants nested within group, and (2) models with a single residual error term versus models with independent residual error terms for each group. Our final models included random intercepts for participants, nested within group, and independent residual errors by group. Models included all available data, whether or not participants completed all sessions. We considered p < 0.05 statistically significant.

## Results

### Participant Characteristics

A total of 22 PwCF and 7 healthy volunteers (HV) consented to participate in this study. Prior to the on-site visit, 3 PwCF withdrew consent. The final sample for analysis was 19 PwCF and 7 HV.

Baseline demographic and anthropometric characteristics are shown in **Table 1**. Both groups were just over 30 years old at enrollment. Just over half of PwCF were male (10/19, 53%), compared with just under half of HV (3/7, 43%). Weight and height were similar between groups. All participants were non-Hispanic and White. Among PwCF, 53% were F508del homozygous and 42% were F508del heterozygous; and all but one (18/19, 95%) were treated with CFTR modulators, predominantly elexacaftor/tezacaftor/ivacaftor.

**Table 1.**
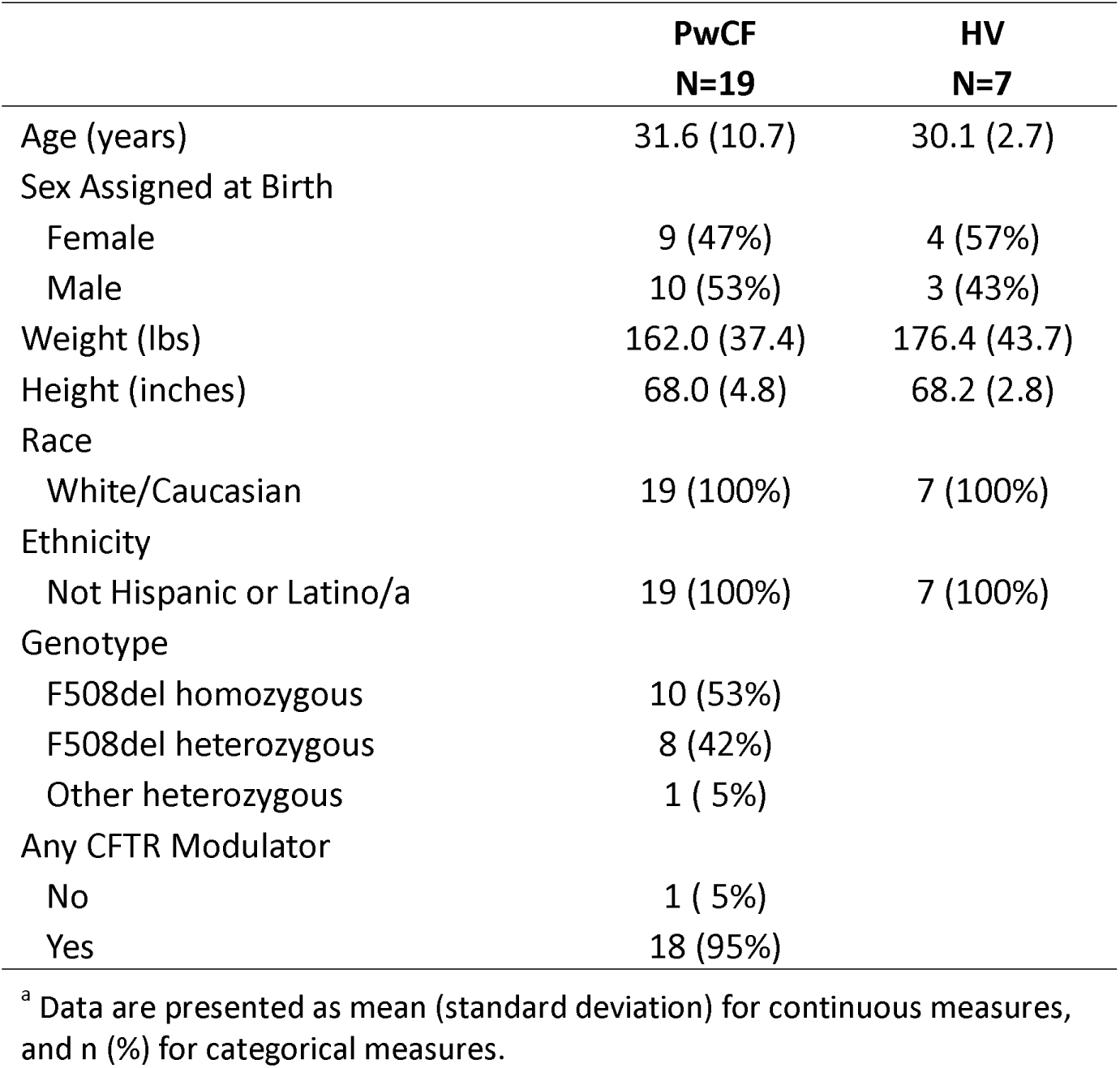
Sample Characteristics among Healthy Volunteers (HV) and Persons with Cystic Fibrosis (PwCF)

|  | **PwCF**<br>**N=19** | **HV**<br>**N=7** |
| --- | --- | --- |
| Age (years) | 31.6 (10.7) | 30.1 (2.7) |
| Sex Assigned at Birth |  |  |
| Female | 9 (47%) | 4 (57%) |
| Male | 10 (53%) | 3 (43%) |
| Weight (lbs) | 162.0 (37.4) | 176.4 (43.7) |
| Height (inches) | 68.0 (4.8) | 68.2 (2.8) |
| Race |  |  |
| White/Caucasian | 19 (100%) | 7 (100%) |
| Ethnicity |  |  |
| Not Hispanic or Latino/a | 19 (100%) | 7 (100%) |
| Genotype |  |  |
| F508del homozygous | 10 (53%) |  |
| F508del heterozygous | 8 (42%) |  |
| Other heterozygous | 1 ( 5%) |  |
| Any CFTR Modulator |  |  |
| No | 1 ( 5%) |  |
| Yes | 18 (95%) |  |

<sup>a</sup> Data are presented as mean (standard deviation) for continuous measures, and n (%) for categorical measures.

### Peak and Laboratory Submaximal-work Exercise in PwCF and HV

**Table 2** summarizes laboratory CPET and submaximal-work (onsite) exercise characteristics by group as well as associated standardized mean differences (SMD), measuring group differences as multiples of pooled standard deviation (SD). Although PwCF and HV were not significantly different in most onsite measures, standardized mean differences between the groups were generally ‘large’ or ‘moderate.’ During symptom-limited CPET, peak wattage was more than 0.9 SDs lower in PwCF than in HV. Maximum heart rate was 0.6 SDs lower in PwCF than in HV. During submaximal-work laboratory exercise (performed at 50–70% of peak wattage), PwCF were more than 0.8 SDs lower than HV in average respiration rate and average wattage. In both CPET and submaximal-work conditions, PwCF were about 0.5 SDs higher in perceived exertion than HV. Although the difference in whole body sweat loss was ‘small’ between groups (0.3 SDs), PwCF lost 0.6 SDs more chloride than HV in onsite exercise sessions.

**Table 2.** Onsite Exercise Characteristics for Healthy Volunteers (HV) and Persons with Cystic Fibrosis (PwCF)

|  | **PwCF**<br>**(N = 19)**<br>**Mean (SD)** | **HV**<br>**(N = 7)**<br>**Mean (SD)** | **SMD** | **(95% CI)** |
| --- | --- | --- | --- | --- |
| **CPET** |  |  |  |  |
| VO2 Max (mL/kg/min) | 30.9 (7.0) | 34.4 (3.0) | -0.54 | (-1.38, 0.32) |
| VO2 Peak Time (minutes) | 9.6 (1.7) | 11.4 (1.7) | -1.02 | (-1.95, -0.08) |
| Peak Wattage | 198.7 (62.1) | 254.3 (49.4) | -0.91 | (-1.78, -0.02) |
| Max HR (bpm) | 171.5 (15.6) | 181.0 (9.5) | -0.64 | (-1.49, 0.22) |
| Max RR (breaths/min) | 42.1 (7.9) | 41.0 (5.8) | 0.14 | (-0.75, 1.02) |
| Maximum Perceived Exertion (1-10) | 7.6 (2.1) | 6.4 (1.5) | 0.55 | (-0.31, 1.40) |
| **Submaximal Exercise** |  |  |  |  |
| Duration (minutes) | 41.2 (8.0) | 36.4 (3.8) | 0.64 | (-0.22, 1.50) |
| Average HR (bpm) | 146.7 (12.9) | 154.7 (19.8) | -0.52 | (-1.37, 0.33) |
| Average RR (breaths/min) | 27.0 (3.9) | 30.3 (4.1) | -0.81 | (-1.72, 0.12) |
| Average Wattage | 96.7 (36.5) | 126.6 (10.4) | -0.90 | (-1.77, -0.02) |
| Perceived Exertion (1-10) | 5.9 (1.2) | 5.3 (0.5) | 0.55 | (-0.31, 1.40) |
| Exercise Volume (bpm * min) | 6022.8 (1206.4) | 5660.9 (1112.4) | 0.30 | (-0.55, 1.14) |
| **Fluid Loss Data** |  |  |  |  |
| Whole Body Sweat Loss (mL) | 367.6 (84.8) | 394.5 (67.2) | -0.32 | (-1.16, 0.52) |
| Total Chloride Loss (mg)<sup>a</sup> | 791.0 (268.5) | 618.7 (275.9) | 0.61 | (-0.28, 1.49) |

SD = Standard deviation. SMD = Standardized mean difference, as measured by Hedge's g.
<sup>a</sup> Onsite chloride loss is missing for four PwCF.

### Remote Exercise Performance versus Laboratory Submaximal-Work (Onsite) Exercise

Among 19 PwCF, 12 had successful heart rate monitoring from all five remote exercise sessions, with 6 of those individuals undergoing successful sweat monitoring in all five sessions as well. Among HV, 5 had successful heart rate monitoring from all five remote sessions, while 3 had successful sweat monitoring from all five sessions. The remainder completed heart rate and sweat monitoring as detailed in Supplemental Table 1. Subsequent analyses utilize all available data.

**Table 3** compares laboratory submaximal-work exercise (onsite) versus remote exercise by group. During remote exercise, PwCF and HV tended to exercise about quarter hour longer but at lower average heart rates (respectively, 14 bpm lower and 19 bpm lower) than onsite exercise. Because participants exercised longer during remote exercise, exercise volume was still higher in both groups during remote exercise sessions (PwCF 1451 bpm*min [95% CI 489, 2412]; HV 1185 bpm*min [95% CI 143, 2227]) than onsite. PwCF perceived their exertion as 0.5 units higher during remote exercise compared with onsite (95% CI 0.05, 1.0); for HV, there was no significant difference. Both groups also lost more whole body sweat fluid during remote exercise: 100.2mL (95% CI42.6, 157.9) for PwCF and 54.0 (95% CI 5.1, 102.8) mL for HV. Total chloride loss was higher during remote exercise for PwCF (149.8 mg, 95% CI 28.6, 271.0), but not for HV.

**Table 3.**
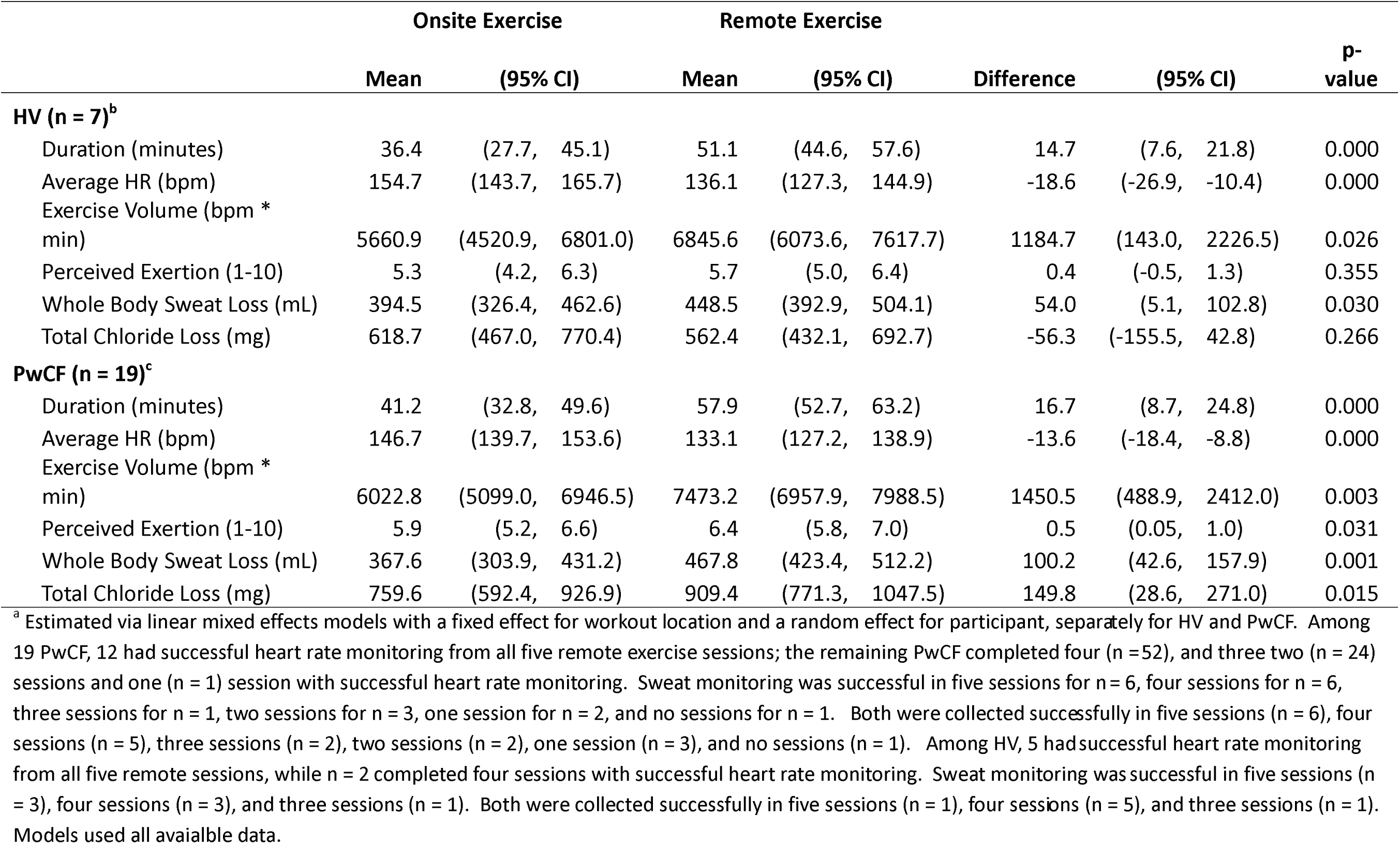
Onsite versus Remote Exercise Characteristics for Healthy Volunteers (HV) and Persons with Cystic Fibrosis (PwCF)

|  | Onsite Exercise |  |  | Remote Exercise |  |  |  |  |  | p-value |
| --- | --- | --- | --- | --- | --- | --- | --- | --- | --- | --- |
|  | Mean | (95% CI) |  | Mean | (95% CI) |  | Difference | (95% CI) |  |  |
| HV (n = 7)<sup>b</sup> |  |  |  |  |  |  |  |  |  |  |
| Duration (minutes) | 36.4 | (27.7, | 45.1) | 51.1 | (44.6, | 57.6) | 14.7 | (7.6, | 21.8) | 0.000 |
| Average HR (bpm) | 154.7 | (143.7, | 165.7) | 136.1 | (127.3, | 144.9) | -18.6 | (-26.9, | -10.4) | 0.000 |
| Exercise Volume (bpm * min) | 5660.9 | (4520.9, | 6801.0) | 6845.6 | (6073.6, | 7617.7) | 1184.7 | (143.0, | 2226.5) | 0.026 |
| Perceived Exertion (1-10) | 5.3 | (4.2, | 6.3) | 5.7 | (5.0, | 6.4) | 0.4 | (-0.5, | 1.3) | 0.355 |
| Whole Body Sweat Loss (mL) | 394.5 | (326.4, | 462.6) | 448.5 | (392.9, | 504.1) | 54.0 | (5.1, | 102.8) | 0.030 |
| Total Chloride Loss (mg) | 618.7 | (467.0, | 770.4) | 562.4 | (432.1, | 692.7) | -56.3 | (-155.5, | 42.8) | 0.266 |
| PwCF (n = 19)<sup>c</sup> |  |  |  |  |  |  |  |  |  |  |
| Duration (minutes) | 41.2 | (32.8, | 49.6) | 57.9 | (52.7, | 63.2) | 16.7 | (8.7, | 24.8) | 0.000 |
| Average HR (bpm) | 146.7 | (139.7, | 153.6) | 133.1 | (127.2, | 138.9) | -13.6 | (-18.4, | -8.8) | 0.000 |
| Exercise Volume (bpm * min) | 6022.8 | (5099.0, | 6946.5) | 7473.2 | (6957.9, | 7988.5) | 1450.5 | (488.9, | 2412.0) | 0.003 |
| Perceived Exertion (1-10) | 5.9 | (5.2, | 6.6) | 6.4 | (5.8, | 7.0) | 0.5 | (0.05, | 1.0) | 0.031 |
| Whole Body Sweat Loss (mL) | 367.6 | (303.9, | 431.2) | 467.8 | (423.4, | 512.2) | 100.2 | (42.6, | 157.9) | 0.001 |
| Total Chloride Loss (mg) | 759.6 | (592.4, | 926.9) | 909.4 | (771.3, | 1047.5) | 149.8 | (28.6, | 271.0) | 0.015 |

<sup>a</sup> Estimated via linear mixed effects models with a fixed effect for workout location and a random effect for participant, separately for HV and PwCF. Among 19 PwCF, 12 had successful heart rate monitoring from all five remote exercise sessions; the remaining PwCF completed four (n = 52), and three two (n = 24) sessions and one (n = 1) session with successful heart rate monitoring. Sweat monitoring was successful in five sessions for n = 6, four sessions for n = 6, three sessions for n = 1, two sessions for n = 3, one session for n = 2, and no sessions for n = 1. Both were collected successfully in five sessions (n = 6), four sessions (n = 5), three sessions (n = 2), two sessions (n = 2), one session (n = 3), and no sessions (n = 1). Among HV, 5 had successful heart rate monitoring from all five remote sessions, while n = 2 completed four sessions with successful heart rate monitoring. Sweat monitoring was successful in five sessions (n = 3), four sessions (n = 3), and three sessions (n = 1). Both were collected successfully in five sessions (n = 1), four sessions (n = 5), and three sessions (n = 1). Models used all available data.

### Comparison of Remote Exercise Characteristics and Fluid Loss between HV and PwCF

During remote exercise sessions, PwCF and HV did not differ significantly in duration, average heart rate, exercise volume, or perceived exertion (**Table 4**). Both groups lost comparable amounts of whole body sweat fluid (mean difference 19.9 mL [95% CI −58.1, 97.8]). However, PwCF lost 917.9 mg total chloride (95% CI 775.3, 1060.6), compared with 560.4 mg chloride (95% CI 443.1, 677.8) among HV, a difference of 357.5 mg (95% CI 172.8, 542.2).

**Table 4.** Differences in Remote Exercise Characteristics Between Healthy Volunteers (HV) and Persons with Cystic Fibrosis (PwCF)

|  | PwCF (N = 19) |  | HV (N = 7) |  | Difference | (95% CI) | p-value |
| --- | --- | --- | --- | --- | --- | --- | --- |
|  | Mean | (95% CI) | Mean | (95% CI) |  |  |  |
| Duration (minutes) | 57.9 | (52.0, 63.9) | 51.1 | (43.7, 58.4) | 6.8 | (-2.6, 16.3) | 0.156 |
| Average HR (bpm) | 132.9 | (126.7, 139.1) | 136.1 | (128.1, 144.1) | -3.2 | (-13.3, 7.0) | 0.539 |
| Exercise Volume (bpm * min) | 7486.0 | (6897.4, 8074.5) | 6847.0 | (6045.6, 7648.5) | 638.9 | (-355.4, 1633.3) | 0.208 |
| Perceived Exertion (1-10) | 6.4 | (5.8, 7.1) | 5.7 | (4.8, 6.6) | 0.7 | (-0.4, 1.8) | 0.195 |
| Whole Body Sweat Loss (mL) | 468.4 | (420.6, 516.2) | 448.6 | (386.9, 510.2) | 19.9 | (-58.1, 97.8) | 0.618 |
| Total Chloride Loss (mg) | 917.9 | (775.3, 1060.6) | 560.4 | (443.1, 677.8) | 357.5 | (172.8, 542.2) | 0.000 |

a Estimated via linear mixed effects models with a fixed effect for diagnosis of CF and independent random effects for participant nested within group (HV vs PwCF), and independent residual errors allowing for different variances for HV and PwCF. Among 19 PwCF, 12 had successful heart rate monitoring from all five remote exercise sessions; the remaining PwCF completed four (n = 52), and three two (n = 24) sessions and one (n = 1) session with successful heart rate monitoring. Sweat monitoring was successful in five sessions for n = 6, four sessions for n = 6, three sessions for n = 1, two sessions for n = 3, one session for n = 2, and no sessions for n = 1. Both were collected successfully in five sessions (n = 6), four sessions (n = 5), three sessions (n = 2), two sessions (n = 2), one session (n = 3), and no sessions (n = 1). Among HV, 5 had successful heart rate monitoring from all five remote sessions, while n = 2 completed four sessions with successful heart rate monitoring. Sweat monitoring was successful in five sessions (n = 3), four sessions (n = 3), and three sessions (n = 1). Both were collected successfully in five sessions (n = 1), four sessions (n = 5), and three sessions (n = 1). Models used all available data.

### Increased Variability in Exercise-Induced Chloride Loss in PwCF Compared to HV

Figure 1 illustrates the relationship between total chloride loss and exercise volume, separately for PwCF and HV. Lines indicate the average relationship between exercise volume and total chloride loss for each group. The slopes show that for every unit increase in exercise volume, participants lost an additional 0.058 (95% CI 0.037, 0.080) mg total chloride, regardless of group. Even after adjusting for exercise volume, PwCF lost an average of 280.94 mg more total chloride than HV (95% CI 85.26 – 476.63) during remote exercise sessions, as illustrated by the vertical distance between the lines. We also evaluated intra-individual variability across remote sessions. From session to session, and after adjusting for both exercise volume and more total chloride loss in PwCF, PwCF had greater variability in total chloride loss (residual standard deviation = 181.52 mg [95% CI143.97, 219.06]) compared with HV (residual standard deviation = 102.07 mg [95% CI 71.18, 132.96]).

**Figure 1.**
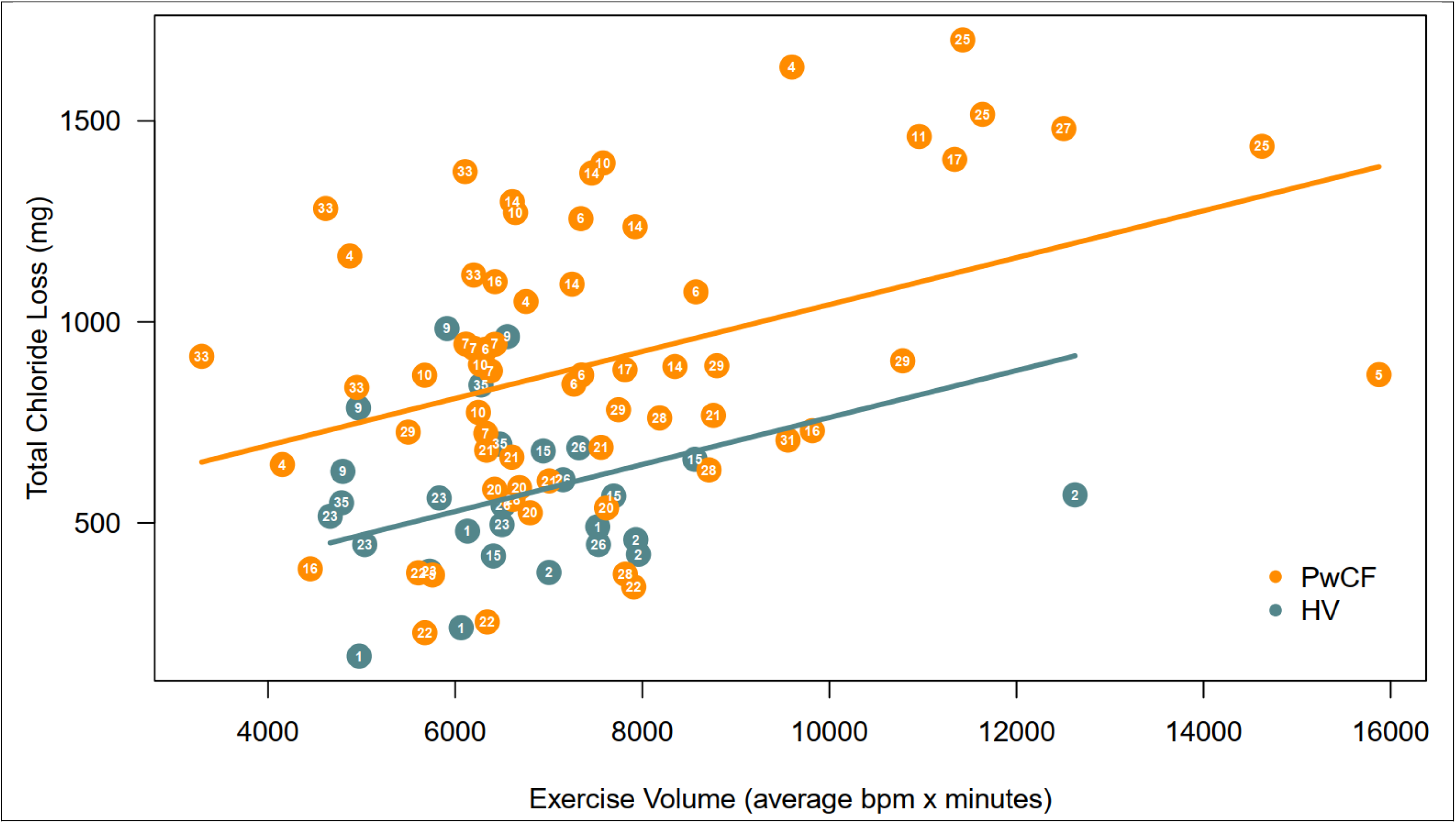
Exercise-associated sweat chloride loss is greater and more variable in people with Cystic Fibrosis compared to healthy volunteers. Each point represents the total sweat chloride (mg) lost during an individual exercise session (y-axis) and exercise volume (x-axis) in people with cystic fibrosis and healthy volunteers. In both groups, total chloride loss increased with greater exercise volume, consistent with higher sweat production and electrolyte loss during more prolonged or intense exercise. Compared with healthy volunteers, people with cystic fibrosis lost greater amounts of chloride for a given exercise volume. Chloride loss was also more variable among people with cystic fibrosis, both across the study population and within individual participants across repeated exercise sessions. These findings demonstrate that exercise-induced chloride loss is influenced by exercise volume but is amplified and more heterogeneous in people with cystic fibrosis.

## Discussion

This study extends the current understanding of exercise physiology in PwCF by integrating wearable microfluidic sweat sensing with supervised and remote exercise testing to quantify real-world fluid and chloride losses. We report several novel observations. First, we establish the feasibility and physiologic utility of directly quantifying exercise-associated chloride loss using a wearable sweat-sensing platform. Second, we demonstrate that PwCF can achieve exercise volumes comparable or even higher than that measured in the exercise laboratory. Third, we show that chloride loss during remote exercise is substantially higher and more variable in PwCF than in HV, despite widespread use of highly effective CFTR modulators. These findings highlight the potential role of sweat chloride monitoring in personalized hydration and electrolyte management and in CFTR biomarker assessment outside clinical settings.

Exercise is recommended as a central component of CF care, with benefits encompassing aerobic capacity, muscle function, bone health, and quality of life.(4–6, 18–20) Prior studies of exercise in CF have primarily focused on peak oxygen uptake, ventilatory limitation, heart rate response, and training effects.(2, 19, 21) Comparatively little work has characterized electrolyte and salt loss during exercise,(22) and most prior sweat studies in CF have relied on pilocarpine iontophoresis under resting, clinic-based conditions.(23–25) Our findings add an important physiologic dimension to the CF exercise literature by linking exercise workload with directly measured sweat fluid and chloride loss under both controlled and free-living conditions. Compared to laboratory sessions, PwCF and HV achieved higher exercise volumes during remote sessions, notably exercising at slightly lower intensity for longer duration. This pattern is consistent with adaptive pacing behavior and supports the concept that meaningful exercise dose in PwCF is not fully captured by peak metrics alone. By pairing exercise volume with quantitative sweat chloride and fluid loss measurements, this study provides a more integrated view of physiologic stress during exercise in CF. In addition, the assessment of heart rate, perceived exertion, and exercise volume between laboratory and remote sessions supports the validity of unsupervised, real-world exercise assessments when coupled with wearable physiologic monitoring. This has implications for both clinical research and decentralized trial design in CF.

A novel observation is the marked variability in exercise-associated chloride loss in PwCF across repeated sessions, exceeding that observed in HV. This variability was evident both between individuals and within the same individual across days, even after accounting for heart-rate–derived exercise volume. This finding is important in the context of CFTR modulator therapy. While modulators substantially reduce sweat chloride on standardized testing, prior work has shown residual heterogeneity in CFTR functional correction.(12) Our results extend these observations by demonstrating that chloride loss during physiologic stress (exercise) remains dynamic and variable in daily life. The mechanisms underlying this variability remain unexplained, but several mechanisms may potentially contribute, including fluctuating CFTR activity, hydration status, environmental conditions, exercise modality, and adherence timing relative to modulator dosing.

From a biologic perspective, the increased variability in chloride loss among PwCF is consistent with partial and variable restoration of CFTR-mediated ductal chloride reabsorption rather than uniform normalization. From a clinical perspective, this variability helps explain why PwCF may have inconsistent symptoms related to salt depletion during exercise or heat exposure despite being on modulator therapy. Interestingly, in healthy individuals, exercise-induced salt loss is flow-rate dependent,(26) which would allow for some baseline variability in chloride loss during free exercise. That we see higher variability in exercise-induced chloride loss in PwCF would suggest that CFTR function and rescue through modulator use also has temporal variability, reinforcing the idea that the observed variability in PwCF is disease-related rather than purely methodological or environmental.

Direct quantification of chloride loss during exercise offers several practical and translational advantages. First, it enables individualized electrolyte replacement strategies. Current guidance for salt supplementation in CF is largely generalized, based on diagnosis and broad activity categories rather than measured losses.(27) Wearable quantification of sweat chloride and volume enables accurate estimation of total salt loss in real time, creating the opportunity for personalized repletion recommendations based on measured rather than assumed need.

Second, it provides a dynamic biomarker of CFTR function under physiologic stress. Standard sweat testing is static and clinic-bound. Exercise-associated sweat chloride measurement captures CFTR-dependent ion transport in a stimulated, real-world state and may be more sensitive to short-term biologic fluctuations. Serial measurements could potentially be used to monitor modulator response, adherence patterns, or pharmacodynamic variability at a higher temporal resolution than periodic clinic testing.

Third, it supports remote and decentralized monitoring models. The feasibility of acquiring interpretable heart rate and sweat chloride and volume data across repeated remote exercise sessions demonstrates that high-quality physiologic and electrolyte data can be collected outside specialized centers. This opens the door to longitudinal home monitoring, integration into digital health platforms, and deployment in geographically distributed populations.

Fourth, it links exercise dose to electrolyte consequences. By pairing chloride loss with exercise volume metrics, clinicians and investigators can move beyond intensity-based prescriptions toward physiology-informed exercise guidance, particularly for patients at higher risk of dehydration or salt depletion. More generally, understanding the electrolyte depletion associated with remote exercise prescriptions of varying intensity and duration could inform general guidelines for PwCF.

This study is a single-center pilot feasibility study with attendant limitations. It has a small sample size and was not powered to detect specific clinical or physiologic effects. Further, many participants did not perform all 5 sessions of remote exercise, reducing data points for assessing variability between exercise sessions. Healthy volunteers were fewer in number and not specifically matched to PwCF. Remote exercise was not specified in the protocol and was less intense than laboratory exercise, contributing to variability, though both cohorts chose their modes, intensity, and duration of exercise. The PwCF were relatively healthy. Most were on highly effective modulators, all were White and non-Hispanic, and 95% had at least one F508del variant, all which limit generalizability. Larger, matched studies are needed to confirm findings and assess clinical utility.

In summary, these findings support the feasibility of a shift from static, clinic-based sweat testing toward contextual, activity-linked sweat phenotyping in CF. For clinicians, quantifying chloride loss during exercise can refine counseling around hydration and salt replacement, particularly for patients engaging in endurance activity, training programs, or exercise in warm environments. For researchers, wearable sweat sensing provides a scalable tool for studying CFTR physiology in daily life and for evaluating interventions that may influence electrolyte homeostasis. More broadly, this work illustrates how wearable biosensing can bridge the gap between molecular correction and real-world physiology in CF. As CF care continues to evolve toward personalized and data-driven management, exercise-linked chloride loss measurement represents a clinically meaningful and mechanistically grounded digital biomarker with immediate translational relevance.

## Author contributions

T.C, R.N. and M.J. conceived the idea, designed the research, performed the data analysis, and wrote the manuscript. A.J.A., R.G., J.A.R., J.A.W., D.S.Y. conceived the idea, designed the research, and wrote the manuscript. S.A.M advised on the study design and wrote the manuscript. L.W and J.S. performed the data analysis and wrote the manuscript. M.G., Z.K., M.C. and A.D. implemented the study protocol. G.J.G. and D.M.C. helped design the protocol and write the manuscript. A.J.A., D.E.W., J.C.W. performed and were involved in the manufacturing of the sensors. A.J.A., J.C.W., D.E.W., performed software design and software validation. D.E.W., J.C.W., A.J.A. assisted in device fabrication and field testing.

## Data Availability

All data produced in the present study are available upon reasonable request to the authors

